# Detection of Biofilm Production of Oral Bacterial Isolates and Its Impact on the Antibiotic Resistance Profile

**DOI:** 10.1101/2025.06.19.25329973

**Authors:** Nesreen Fadel Al-Sanabani, Hassan Abdulwahab Al-Shamahy

## Abstract

**Objectives:** The aim of this study was to investigate the potential association between the formation of oral bacterial biofilms and the occurrence of antibiotic resistance among 294 oral bacterial isolates.

**Study design:** A total of 100; 25-65year old patients were chosen. Buccal mucosa swabs were collected and cultured in appropriate media, then bacteria were isolated and identified. Then 294 bacterial isolates were assessed for biofilm production by the phenotypic method, i.e., the tissue culture plate method (TCPM). Finally, antibiotic susceptibility patterns of the 294 isolates were done by the Kirby-Bauer disc diffusion method for 7 β-lactam antibiotics (ampicillin, penicillin, amoxicillin, cephalothin, oxacillin, cloxacillin, and cefoxitin) and 7 non-β-lactam antibiotics (tetracycline, co-trimoxazole, ciprofloxacin, clindamycin, erythromycin, lincomycin, and vancomycin).

**Results:** When isolates were exposed to biofilm detection by the TCP method, 9 (3.1%) showed high biofilm formation capacity, 213 (72.4%) showed moderate biofilm formation capacity, while 72 (24.5%) showed weak/no biofilm formation. The isolated bacterial biofilms positively showed that the bacterial isolates that showed high and moderate biofilm formation capacity have a higher rate of resistance to most antibiotics with significant difference (p < 0.0001) than weak/no biofilm formation.

**Conclusion:** The present study demonstrates that aerobic bacteria are still the major bacteria isolates from the oral cavity. Antibiotic resistance in the oral bacterial isolates was found to be associated with bacterial biofilm formation.

## INTRODUCTION

The capacity to form biofilms is a feature shared by all microorganisms. In biofilms, an extracellular matrix formed by the cells themselves binds the microbial communities together. Bacterial species employ diverse mechanisms to create biofilms; these mechanisms often depend on the environment in which they are found as well as strain-specific characteristics. Antonie van Leeuwenhoek first saw “animalcules” on his own teeth in the 17th century. The “bottle effect” was first noted in marine microorganisms in 1940 [1]. This demonstrated that bacteria proliferate more abundantly on surfaces. Then, in 1943, Zobell created biofilms and discovered that the amount of bacteria on surfaces was higher than that of the surrounding saltwater [2].

Because of their increased resistance to antibiotics and disinfectants, bacterial biofilms are a major contributing factor to chronic infections. They can also interfere with phagocytosis and other immune system functions [3]. As a result, microorganisms within biofilms become less susceptible to antimicrobial agents, posing significant clinical challenges in the field of therapeutics [4]. However, the development of biofilm, the composition of extracellular matrix (ECM), including proteins, lipids, water, glycolipids, polysaccharides, extracellular DNA, membrane vesicles, and extracellular RNA, and the architecture of biofilm—that is, the biomass and spatial organization within the biofilm—are the main factors influencing antibiotic tolerance [5–8]. Most oral biofilms are made up of several different bacterial strains. Over 700 distinct bacterial species have been shown to be present in tooth plaque recently [9].

Biofilms exhibit both antibiotic tolerance and resistance. Through genetic mutation or the acquisition of foreign genetic material encoding resistance determinants through horizontal gene transfer (HGT) in biofilm EPS, microorganisms evolve defense mechanisms against antimicrobials. Antimicrobial resistance (AMR) is caused by several mechanisms, including decreased permeability or access to antimicrobials, modifications to targets, modification of antimicrobial targets, and enzymatic destruction of the antimicrobials by hydrolysis or chemical change. While antibiotics are effective against bacteria, yet the bacteria develop resistance to them, antibiotic resistance (ABR) is a subset of antimicrobial resistance (AMR) [10,11]. The current investigation set out to determine if oral bacterial isolates produced biofilms and whether this was associated with the antibiotic resistance profiles of the isolates from bacteria that had colonized the mouth mucosa.

## MATERIALS AND METHODS

Participant recruitment took place between June 1, 2022 and July 15, 2022. Two hundred ninety-four (294) bacteria isolated from the mouths of 100 patients attending dental clinics at the Faculty of Dentistry at Sana’a University were examined for the development of biofilms. The antibiotic susceptibility of the isolated bacteria was then phenotypically determined using standard techniques according to the 2015 standards of the Clinical and Laboratory Standards Institute (CLSI) [12,13]. Oral bacteria that produce biofilms have been identified using the tissue culture plate (TCP) method and tested the association of antibiotic resistance with biofilm production.

### Antibiogram

The disc diffusion method was used to determine the antibiotic susceptibility profile. The inoculums were modified to correspond to 0.5 McFarland standards of turbidity, then swabbed on Brain heart infusion agar and left to dry for 10 minutes [12–14]. The susceptibility to non-β-lactam antibiotics, including, erythromycin (15 µg), gentamicin (10 µg), amikacin (30 µg), ciprofloxacin (5 µg), clindamycin (2 µg), and vancomycin (5 µg), tetracycline (30 µg), co-trimoxazole (15 µg), and β-lactam antibiotics, including ampicillin (25µg), penicillin (10 µg), oxacillin (15 µg), Cefoxitin (30 µg), and Amoxicillin-Clavulanic Acid (25 µg) were then assessed using antibiogram profiling (Oxoid, UK). After 24 hours of aerobic incubation at 37°C, the inhibition zone was determined. Each antibiotic’s experiments were carried out three times. Clinical and Laboratory Standards Institute (CLSI) methods were used to analyze the results [12–14].

### Biofilm production detection

Tissue culture/microtiter plate method (TCP) was used to identify biofilm [15]. After being inoculated with 2mL of BHI broth, the bacterial isolates from fresh agar plates were cultured for 24 hours at 37°C. Following a 1:40 dilution with fresh medium (BHI broth supplemented with 1% glucose), 200μL of the sample was added to each individual micro titration plate, and the plates were incubated for an additional 24 hours at 37°C. After lightly tapping the contents, free floating sessile bacteria were eliminated by repeatedly rinsing it with phosphate-buffered saline (pH 7.2). For ten to fifteen minutes, adhering bacteria that produced biofilm were fixed with sodium acetate (2% w/v) and stained with crystal violet (0.1% w/v). After removing the unbound crystal violet solution in three PBS washes, the plate was set aside to dry. In order to release the dye, 200μL of 95% ethanol was added to each well, and an optical density (OD) was measured at 630 nm was taken. Each test strain’s OD value as well as that of the negative control were computed, and OD cutoff values (ODc) were evaluated in accordance with earlier instructions [15].

### Statistical Analysis

Version 7 of EPI-INFO statistical software was used to analyze the data. By computing the difference, 95% CI, and p-value of the antibiotic resistance for each tested antibiotic with level of biofilm production, statistical analysis was carried out to take into account the degree of biofilm production of 294 bacterial isolates with the degree of drug resistance.

### Ethical Consideration

On August 21, 2022, the Medical Ethics and Research Committee of Sana’a University’s Faculty of Medicine and Health Sciences granted ethical approval for this study (Approval No. 217). The ethical rules established by the review committee were consistently followed, and all participants provided signed informed consent.

## RESULTS

Table 1 shows the interpretation of biofilm production by the tested bacteria based on the optical density values of the tissue culture plate method for average biofilm production with an OD values. Nine bacteria (3.1%) showed a no ability to produce biofilms (OD < 0.17), and 21.4% of the tested bacteria showed a low capacity to produce biofilms (OD = 0.17–0.34). Most of the orally isolated bacteria showed moderate production (OD = 0.35-0.68), which amounted to 72.4%, while only 3.1% of the tested bacteria showed strong production for biofilm production (OD > 0.68) (Table 1). Table 2 shows the detection of biofilms by the TCP method among 294 different bacterial species isolated in the oral cavity. Most S. *aureus* strains showed moderate and strong biofilm production (93.8% and 4.2%, respectively), while the other 64.3% of *coagulase-negative staphylococci* strains showed only moderate positive biofilm production. Considering the *viridans* group *streptococci*, *S*. *mutans* had 86.6% strains with moderate biofilm production and 1.2% with strong biofilm production, while other *streptococci* had less biofilm production capacity (Table 2). For *Neisseria* species, 72% showed a moderate level of biofilm production.

**Table 1:** Interpretation of biofilm production by bacterial isolates based on optical density values of tissue culture plate method Average value of OD* Biofilm production.

| OD value | N (%) |
| --- | --- |
| <0.17 Negative | 9 (3.1) |
| 0.17-0.34 Weak positive | 63 (21.4) |
| 0.35-0.68 Moderate positive | 213 (72.4) |
| >0.68 Strong positive | 9 (3.1) |
| Total | 294 (100) |

**Table 2:** Biofilm detection by TCA method among different species of oral cavity isolated Bacteria n=294.

| Bacteria | Biofilm production by TCA |  |  |  |
| --- | --- | --- | --- | --- |
|  | Negative No (%) | Weak No (%) | moderate No (%) | Strong No (%) |
| *S. aureus* n=48 | 0 (0.0) | 1(2.1) | 45 (93.8) | 2 (4.2) |
| Coagulase-negative n=14 | 1(7.1) | 4 (28.6) | 9 (64.3) | 0 (0.0) |
| ***Streptococci*** |  |  |  |  |
| *S. pyogenes* n=3 | 1 (33.3) | 1 (33.3) | 1 (33.3) | 0 (0.0) |
| *S.mitior* n=18 | 0 (0.0) | 7 (38.9) | 11 (61.1) | 0 (0.0) |
| *S.sanguis* n=17 | 1 (5.9) | 9 (52.9) | 6 (35.3) | 1 (5.9) |
| *S.mutans* n=82 | 0 (0.0) | 10 (12.2) | 71 (86.6) | 1 (1.2) |
| *S.salivarius* n=25 | 1 (4) | 7 (28) | 17 (68) | 0 (0.0) |
| *S.milleri* n=4 | 1 (25) | 1 (25) | 2 (25) | 0 (0.0) |
| *Neisseria species* n=50 | 1(2) | 13 (26) | 36 (72) | 0 (0.0) |
| *Haemophilus influenzae* n=1 | 0 (0.0) | 0 (0.0) | 0 (0.0) | 1 (100) |
| *H.parainfluenzae* n=12 | 0 (0.0) | 5 (41.7) | 7 (58.3) | 0 (0.0) |
| ***Enterobacteriaceae spp.*** |  |  |  |  |
| *Escherichia coli* n=7 | 1 (14.3) | 1 (14.3) | 5 (71.4) | 0 (0.0) |
| *Klebsiella pneumoniae* n=4 | 1 (25) | 0 (0.0) | 2 (50) | 1 (25) |
| *Morganella morganii* n=1 | 0 (0.0) | 1 (100) | 0 (0.0) | 0 (0.0) |
| *Citrobacter freundii* n=2 | 0 (0.0) | 0 (0.0) | 1 (50) | 1 (50) |
| *Pseudomonas aeruginosa* n=3 | 1 (33.3) | 1(33.3) | 0 (0.0) | 1 (33.3) |
| *Proteus species* n=2 | 0 (0.0) | 1 (50) | 0 (0.0) | 1 (50) |
| *Enterobacter species* n=1 | 0 (0.0) | 1 (100) | 0 (0.0) | 0 (0.0) |
| Total n=294 | 9 (3.1) | 63 (21.4) | 213 (72.4) | 9 (3.1) |

The relationship between *S. aureus* biofilm development and antibiotic resistance in isolates from patient buccal mucosa is displayed in Table 3. For example, the difference in resistance to ampicillin was 95.1%, indicating that biofilm-producing strains (moderate/strong) are resistant to ampicillin 95.1% when compared with negative/weak strains. This rate ranges between 16.5 and 98.2%, and this result is highly statistically significant, with p<0.0001. In conclusion the rate of drug resistance for S. *aureus* against the antibiotics tested was higher in moderate/strong biofilm-producing strains than in negative/weak biofilm-producing strains. Also for other antibiotics, the rate of drug resistance against the specific antibiotic tested was higher in moderate/strong biofilm-producing strains than in negative/weak biofilm-producing strains. The association between *Coagulase-negative staphylococcus* biofilm growth and antibiotic resistance in isolates from patient buccal mucosa is presented in Table 4. For instance, the difference in tetracycline resistance was 77.8%, meaning that biofilm-producing germs (strong/moderate) are 77.8% more resistant to ampicillin than negative/weak strains. With a *p*-value of 0.005, this rate falls between 23.4 and 91.2%, and the outcome is statistically significant. In conclusion, moderate/strong biofilm-producing strains of *Coagulase-negative staphylococcus* exhibited a higher rate of drug resistance against the tested antibiotics than did negative/weak biofilm-producing strains. In addition, for other antibiotics, the rate of drug resistance was higher in *Coagulase-negative staphylococcus* strains that produced moderate or strong biofilms compared to strains that produced weak or negative biofilms.

**Table 3:** Association of biofilm formation and Antibiotic resistant of *S.aureus* isolated from buccal mucosa of patients n=48 isolates.

|  | Total n=48 | Negative/weak<br>N=1 | Moderate/strong<br>N=47 | DF (95% CI) | p value |
| --- | --- | --- | --- | --- | --- |
| Antibiotic name | Resistance<br>N (%) | Resistance<br>N (%) | Resistance<br>N (%) |  |  |
| Ampicillin | 46 (95.8) | 1(2.2) | 45 (97.8) | 95.1(16.5-98.2) | <0.0001 |
| Tetracycline | 48 (100) | 1 (2.1) | 47 (97.9) | 87.4(11-94.8) | 0.001 |
| Erythromycin | 16 (33.3) | 1 (6.3) | 15 (93.7) | 100(20.3-100) | <0.0001 |
| Cephalothin | 45 (93.8) | 0 (0.0) | 45 (100) | 100(20.3-100) | <0.0001 |
| Co-trimoxazole | 22 (45.8) | 0 (0.0) | 22 (100) | 100(19.3-100) | <0.0001 |
| Amoxicillin-Clavulanic Acid | 26 (54.2) | 0 (0.0) | 26 (100) | 100(19.6-100) | <0.0001 |
| Gentamicin | 5 (10.4) | 0 (0.0) | 5 (100) | 100(9.5-100) | 0.025 |
| Oxacillin | 33 (68.7) | 0 (0.0) | 33(100) | 100(19.9-100) | <0.0001 |
| Penicillin | 36 (75) | 0 (0.0) | 36 (100) | 100(19.9-100) | <0.0001 |
| Ciprofloxacin | 36 (75) | 0 (0.0) | 36 (100) | 100(20.1-100) | <0.0001 |
| Cloxacillin | 22 (45.8) | 0 (0.0) | 22 (100) | 100(19.3-100) | <0.0001 |
| Cefoxitin | 33 (68.7) | 0 (0.0) | 33(100) | 100(19.9-100) | <0.0001 |
| Amikacin | 0 (0) | 0 (0.0) | 0 (100) | - | - |
| Clindamycin | 0 (0) | 0 (0.0) | 0 (100) | - | - |
| Vancomycin | 0 (0) | 0 (0.0) | 0 (100) | - | - |

DF=difference

**Table 4:** Association of biofilm formation and Antibiotic resistant of *Coagulase-negative staphylococcus* isolated from buccal mucosa n=14 isolates.

|  | Resistant | Biofilm production |  | DF (95% CI) | p value |
| --- | --- | --- | --- | --- | --- |
| Antibiotic name | Total<br>N (%) | Negative/weak<br>N=5 | Moderate/strong<br>N=9 |  |  |
| Tetracycline | 9 (64.3) | 1(11.1) | 8 (88.9) | 77.8(23.4-91.2) | 0.005 |
| Erythromycin | 5(35.7) | 1 (20) | 4 (80) | 60(6.4-81.9) | 0.03 |
| Co-trimoxazole | 6 (42.9) | 1 (16.7) | 5 (83.3) | 66.6(12.6-85.6) | 0.01 |
| Amoxicillin-Clavulanic Acid | 5(35.7) | 0 (0.0) | 5 (100) | 100(47.2-100) | 0.0003 |
| Gentamicin | 4 (28.6) | 0 (0.0) | 4 (100) | 100(42.6-100) | 0.0005 |
| Oxacillin | 3 (21.4) | 0 (0.0) | 3 (100) | 100(42.6-100) | 0.0005 |
| Ciprofloxacin | 2 (14.3) | 0 (0.0) | 2 (100) | 100(42.6-100) | 0.0005 |
| Cloxacillin | 2 (14.3) | 0 (0.0) | 2 (100) | 100(42.6-100) | 0.0005 |
| Cefoxitin | 2 (14.3) | 0 (0.0) | 2 (100) | 100(42.6-100) | 0.0005 |
| Amikacin | 0(0.0) | 0(0.0) | 0(0.0) | - | - |
| Clindamycin | 0(0.0) | 0(0.0) | 0(0.0) | - | - |
| Vancomycin | 0(0.0) | 0(0.0) | 0(0.0) | - | - |

The association between *S.mutans* biofilm growth and antibiotic resistance in isolates from patient buccal mucosa is presented in Table 5. For instance, the difference in tetracycline resistance was 75.6%, meaning that biofilm-producing germs (strong/moderate) are 75.6% more resistant to tetracycline than negative/weak strains, with a p-value of <0.0001, this rate falls between 43.4 and 86.8%, and the outcome is statistically significant (p<0.0001). In conclusion, moderate/strong biofilm-producing strains of *S.mutans* exhibited a higher rate of drug resistance against the tested antibiotics than did negative/weak biofilm-producing strains. In addition, for other antibiotics, the rate of drug resistance was higher in *S.mutans* strains that produced moderate or strong biofilms compared to strains that produced weak or negative biofilms. The association between *S. mitis* biofilm growth and antibiotic resistance in isolates from patient buccal mucosa is presented in Table 6. For instance, the difference in tetracycline resistance was 100%, meaning that biofilm-producing germs (strong/moderate) are 100% resistant to tetracycline than negative/weak strains, with a p-value of <0.0001, this rate falls between 56.1 and 100%, and the outcome is statistically significant (p<0.0001). In conclusion, moderate/strong biofilm-producing strains of *S. mitis* exhibited a higher rate of drug resistance against the tested antibiotics than did negative/weak biofilm-producing strains. In addition, for other antibiotics, the rate of drug resistance was higher in *S. mitis* strains that produced moderate or strong biofilms compared to strains that produced weak or negative biofilms. The association between *S. sanguinis* biofilm growth and antibiotic resistance in isolates from patient buccal mucosa is presented in Table 7. For instance, the difference in tetracycline resistance was 71.4%, meaning that biofilm-producing germs (strong/moderate) are 71.4% resistant to tetracycline than negative/weak strains, with a p-value of 0.0046, this rate falls between 23.2 and 100%, and the outcome is statistically significant (p=0.0046). In conclusion, moderate/strong biofilm-producing strains of *S. sanguinis* exhibited a higher rate of drug resistance against the tested antibiotics than did negative/weak biofilm-producing strains. In addition, for other antibiotics, the rate of drug resistance was higher in *S. sanguinis* strains that produced moderate or strong biofilms compared to strains that produced weak or negative biofilms. The association between *S. salivarius* biofilm growth and antibiotic resistance in isolates from patient buccal mucosa is presented in Table 8. For instance, the difference in tetracycline resistance was 77.8%, meaning that biofilm-producing germs (strong/moderate) are 77.8% resistant to tetracycline than negative/weak strains, with a p-value of 0.0002, this rate falls between 36.6 and 90.1%, and the outcome is statistically significant (p = 0.0002). In conclusion, moderate/strong biofilm-producing strains of *S. salivarius* exhibited a higher rate of drug resistance against the tested antibiotics than did negative/weak biofilm-producing strains. In addition, for other antibiotics, the rate of drug resistance was higher in *S. salivarius* strains that produced moderate or strong biofilms compared to strains that produced weak or negative biofilms.

**Table 5:** Association of biofilm formation and Antibiotic resistant of *S.mutans* isolated from buccal mucosa n= 82 isolates.

|  | Resistance | Biofilm formation |  | DF (95% CI) | p value |
| --- | --- | --- | --- | --- | --- |
| Antibiotic name | Total N (%) | Negative/weak N=10 | Moderate/strong N=72 |  |  |
| Tetracycline | 82 (100) | 10 (12.2) | 72 (87.8) | 75.6 (43.4-86.8) | <0.0001 |
| Erythromycin | 0 (0.0) | 0 (0.0) | 0 (0.0) | - | - |
| Ampicillin | 79 (96.3) | 7 (8.9) | 72 (91.1) | 82.2 (50.6-90.9) | <0.0001 |
| Cephalothin | 40 (48.8) | 1 (2.5) | 39 (97.5) | 95 (65.5 -97.9) | <0.0001 |
| Co-trimoxazole | 10 (12.2) | 1 (10) | 9 (90) | 80 (48.3-89.6) | <0.0001 |
| Amoxicillin-Clavulanic Acid | 3 (3.7) | 0 (0.0) | 3 (100) | 100 (71.7 - 100) | <0.0001 |
| Gentamicin | 6 (7.3) | 0 (0.0) | 6 (100) | 100 (71.7 - 100) | <0.0001 |
| Oxacillin | 5 (6.1) | 0 (0.0) | 5 (100) | 100 (71.7 - 100) | <0.0001 |
| Penicillin | 0 (0.0) | 0 (0.0) | 0 (0.0) | - | - |
| Ciprofloxacin | 4 (4.9) | 0 (0.0) | 4 (100) | 100 (71.7 - 100) | <0.0001 |
| Cloxacillin | 5 (6.1) | 0 (0.0) | 5 (100) | 100 (71.7 - 100) | <0.0001 |
| Cefoxitin | 7 (8.5) | 0 (0.0) | 7 (100) | 100 (71.7 - 100) | <0.0001 |
| Amikacin | 0 (0.0) | 0 (0.0) | 0 (0.0) | - | - |
| Clindamycin | 16 (19.5) | 0 (0.0) | 16 (100) | 100 (71.7 - 100) | <0.0001 |
| Vancomycin | 0 (0.0) | 0 (0.0) | 0 (0.0) | - | - |

**Table 6:** Association of biofilm formation and Antibiotic resistant of *S.mitior* isolated from buccal mucosa n= 18 isolates.

|  |  | Biofilm formation |  | DF (95% CI) | p value |
| --- | --- | --- | --- | --- | --- |
| Antibiotic name | Resistance N (%) | Negative/weak N=7 | Moderate/strong N=11 |  |  |
| Tetracycline | 8 (44.4) | 0 (0.0) | 8 (100) | 100 (56.1-100) | <0.0001 |
| Erythromycin | 2 (11.1) | 0 (0.0) | 2 (100) | 100 (25.7-100) | 0.004 |
| Co-trimoxazole | 3 (16.7) | 0 (0.0) | 3 (100) | 100 (33.6-100) | 0.002 |
| Amoxicillin-Clavulanic Acid | 6 (33.3) | 0 (0.0) | 6 (100) | 100 (47.3-100) | 0.0005 |
| Gentamicin | 2 (11.1) | 0 (0.0) | 2 (100) | 100 (25.7-100) | 0.004 |
| Oxacillin | 0 (0.0) | 0 (0.0) | 0 (0.0) | - | - |
| Ciprofloxacin | 0 (0.0) | 0 (0.0) | 0 (0.0) | - | - |
| Cloxacillin | 0 (0.0) | 0 (0.0) | 0 (0.0) | - | - |
| Cefoxitin | 0 (0.0) | 0 (0.0) | 0 (0.0) | - | - |
| Amikacin | 0 (0.0) | 0 (0.0) | 0 (0.0) | - | - |
| Clindamycin | 0 (0.0) | 0 (0.0) | 0 (0.0) | - | - |
| Vancomycin | 0 (0.0) | 0 (0.0) | 0 (0.0) | - | - |

**Table 7:** Association of biofilm formation and Antibiotic resistant of *S.sanguis* isolated from buccal mucosa n= 17 isolates.

| Antibiotic name | Resistance N (%) | Biofilm formation |  | DF (95% CI) | p value |
| --- | --- | --- | --- | --- | --- |
|  |  | Negative/weak N=10 | Moderate/strong N=7 |  |  |
| Tetracycline | 7 (41.2) | 1 (14.3) | 6 (85.7) | 71.4 (23.2-100) | 0.0046 |
| Erythromycin | 3 (17.6) | 0 (0.0) | 3 (100) | 100 (37.4-100) | 0.0005 |
| Co-trimoxazole | 3 (17.6) | 0 (0.0) | 3 (100) | 100 (37.4-100) | 0.0005 |
| Amoxicillin-Clavulanic Acid | 2 (11.8) | 0 (0.0) | 2 (100) | 100 (28.6-100) | 0.0009 |
| Gentamicin | 0 (0.0) | 0 (0.0) | 0 (0.0) | - | - |
| Oxacillin | 0 (0.0) | 0 (0.0) | 0 (0.0) | - | - |
| Ciprofloxacin | 0 (0.0) | 0 (0.0) | 0 (0.0) | - | - |
| Cloxacillin | 2 (11.8) | 0 (0.0) | 2 (100) | 100 (28.6-100) | 0.0009 |
| Cefoxitin | 0 (0.0) | 0 (0.0) | 0 (0.0) | - | - |
| Amikacin | 0 (0.0) | 0 (0.0) | 0 (0.0) | - | - |
| Clindamycin | 0 (0.0) | 0 (0.0) | 0 (0.0) | - | - |
| Vancomycin | 0 (0.0) | 0 (0.0) | 0 (0.0) | - | - |

**Table 8:** Association of biofilm formation and Antibiotic resistant of *S.salivarius* isolated from buccal mucosa n= 25 isolates.

| Antibiotic name | Resistance N (%) | Negative/weak N=8 | Moderate/strong N=17 | DF (95% CI) | p value |
| --- | --- | --- | --- | --- | --- |
| Tetracycline | 9 (36) | 1 (11.1) | 8 (88.9) | 77.8(36.6-90.1) | 0.0002 |
| Erythromycin | 5 (20) | 0 (0.00) | 5 (100) | 100 (52-100) | <0.0001 |
| Co-trimoxazole | 6 (24) | 1 (16.7) | 5 (83.3) | 66.6 (24-83.6) | 0.0017 |
| Amoxicillin-Clavulanic Acid | 4 (16) | 0 (0.0) | 4 (100) | 100 (26.6-100) | 0.0027 |
| Gentamicin | 2 (8) | 0 (0.0) | 2 (100) | 100 (28.6-100) | 0.0009 |
| Oxacillin | 4 (16) | 0 (0.0) | 4 (100) | 100 (26.6-100) | 0.0027 |
| Ciprofloxacin | 0 (0.0) | 0 (0.0) | 0 (0.0) | - | - |
| Cloxacillin | 4 (16) | 0 (0.0) | 4 (100) | 100 (26.6-100) | 0.0027 |
| Cefoxitin | 0 (0.0) | 0 (0.0) | 0 (0.0) | - | - |
| Amikacin | 0 (0.0) | 0 (0.0) | 0 (0.0) | - | - |
| Clindamycin | 0 (0.0) | 0 (0.0) | 0 (0.0) | - | - |
| Vancomycin | 0 (0.0) | 0 (0.0) | 0 (0.0) | - | - |

The association between *Neisseria species* biofilm growth and antibiotic resistance in isolates from patient buccal mucosa is presented in Table 9. For instance, the difference in tetracycline resistance was 80%, meaning that biofilm-producing bacteria (strong/moderate) are 80% resistant to tetracycline than negative/weak strains, with p < 0.0001, this rate falls between 51.3 and 89.9%, and the outcome is statistically significant (p < 0.0001). In conclusion, moderate/strong biofilm-producing strains of *Neisseria species* exhibited a higher rate of drug resistance against the tested antibiotics than did negative/weak biofilm-producing strains. In addition, for other antibiotics, the rate of drug resistance was higher in *Neisseria species* strains that produced moderate or strong biofilms compared to strains that produced weak or negative biofilms. The association between *H. parainfluenzae* biofilm growth and antibiotic resistance in isolates from patient buccal mucosa is presented in Table 10. For instance, the difference in tetracycline resistance was 100%, indicating that biofilm-producing strains (strong/moderate) are 100% resistant to tetracycline than negative/weak strains, with a p-value of 0.0047, with values ranging from 34.5 and 100%, showing statistical significance (p = 0.0047). In conclusion, moderate/strong biofilm-producing strains of *H. parainfluenzae* exhibited a higher rate of drug resistance against the tested antibiotics than did negative/weak biofilm-producing strains. In addition, for other antibiotics, the rate of drug resistance was higher in *H. parainfluenzae* strains that produced moderate or strong biofilms compared to strains that produced weak or negative biofilms.

**Table 9:**
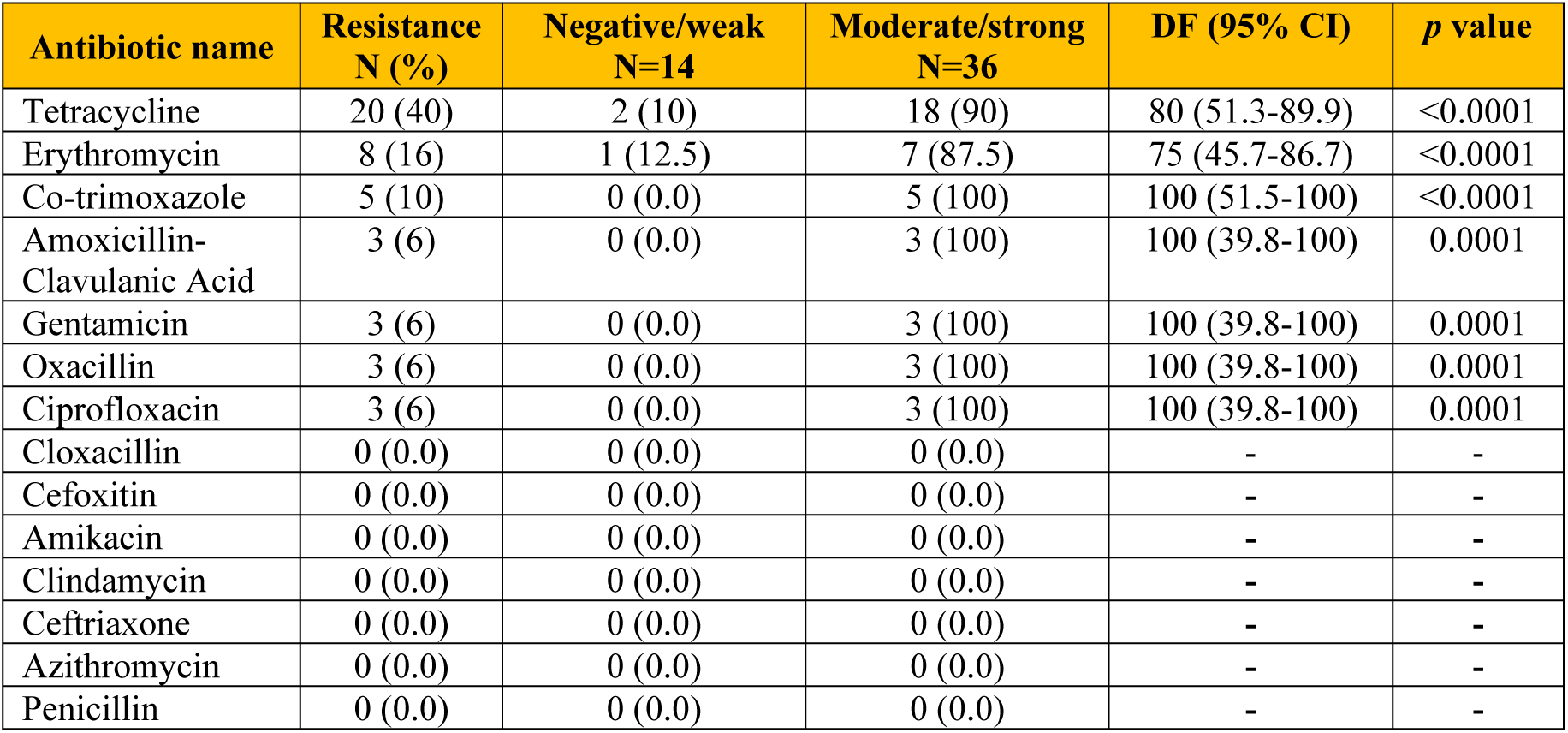
Association of biofilm formation and Antibiotic resistant of *Neisseria species* isolated from buccal mucosa n= 50 isolates.

**Table 10:** Association of biofilm formation and Antibiotic resistant of *H.parainfluenzae* isolated from buccal mucosa of patients n= 12 isolates.

| Antibiotic name | Resistance<br>N (%)<br>N=12 | Negative/weak<br>N=5 | Moderate/strong<br>N=7 | DF (95% CI) | p value |
| --- | --- | --- | --- | --- | --- |
| Tetracycline | 4 (33.3) | 0 (0.0) | 4 (100) | 100 (34.5 -100) | 0.0047 |
| Erythromycin | 1(8.3) | 0 (0.0) | 1(100) | 100 (9.5-100) | 0.025 |
| Co-trimoxazole | 1(8.3) | 0 (0.0) | 1 (100) | 100 (51.5-100) | <0.0001 |
| Amoxicillin-Clavulanic Acid | 4 (33.3) | 0 (0.0) | 4 (100) | 100 (34.5-100) | 0.0047 |
| Gentamicin | 2 (16.7) | 0 (0.0) | 2 (100) | 100 (21.1-100) | 0.01 |
| Oxacillin | 2 (16.7) | 0 (0.0) | 2 (100) | 100 (21.1-100) | 0.01 |
| Ciprofloxacin | 0 (0.0) | 0 (0.0) | 0 (0.0) | - | - |
| Cloxacillin | 0 (0.0) | 0 (0.0) | 0 (0.0) | - | - |
| Cefoxitin | 0 (0.0) | 0 (0.0) | 0 (0.0) | - | - |
| Amikacin | 0 (0.0) | 0 (0.0) | 0 (0.0) | - | - |
| Clindamycin | 0 (0.0) | 0 (0.0) | 0 (0.0) | - | - |
| Vancomycin | 0 (0.0) | 0 (0.0) | 0 (0.0) | - | - |

## DISCUSSION

Finding a relationship between the oral cavity pathogenic bacteria’s ability to form biofilms and its susceptibility to antimicrobial agents, as demonstrated by the patient samples, was a critical goal. When comparing biofilm producers to non-biofilm producers, there was a significant increase in antibiotic resistance. These findings align with studies by Pramodhini et al. [16], Maqbool et al. [17], Tayal et al. [18], Alhasani et al. [10], and Alhasani et al. [11], which found that biofilm producers had significantly higher antibiotic resistance than non-biofilm producers. The level of antibiotic resistance in biofilm producers may be higher than in non-biofilm isolates in the current study and earlier investigations. This could be due to biofilm formation, which have a lower rate of bacterial growth and long-term organism persistence in a variety of conditions. Additionally, as noted by Abdagire et al., [19], close cellular proximity inside a biofilm might promote plasmid exchange and hence accelerate the spread of antibiotic resistance. Amikacin, clindamycin, and vancomycin demonstrated the most effective antibiotics against biofilm-producing Gram positive isolates from the oral cavity in the current study. These findings are consistent with those previously reported by Alhasani et al. [10,11].

In the current study, most of the orally isolated bacteria showed moderate biofilm production (OD = 0.35-0.68), which amounted to 72.4%, while only 3.1% of the tested bacteria showed strong biofilm production (Table 1), in which *S*. *aureus* strains had the highest rate of moderate and strong biofilm production (93.8% and 4.2%, respectively), and the difference in resistance to ampicillin was 95.1%, indicating that biofilm-producing strains (moderate/strong) are resistant to ampicillin 95.1% when compared with negative/weak strains. This rate ranges between 16.5 and 98.2%, and this result is highly statistically significant, with p < 0.0001. These findings reflect the fact that S. *aureus* is one of the most prevalent multidrug-resistant pathogens that is linked to high rates of mortality and morbidity worldwide. Additionally, because AMR *S. aureus* forms biofilms, managing neonatal sepsis has become more difficult. Failure of first- and second-generation antibiotics unavoidably pushes the creation of future generation antimicrobials, requiring significant resources. Several researchers [20–26] have reported biofilm-forming MDR *S. aureus* to be resistant to vancomycin, a second-line medication, and linezolid, a last-resort medication; however, no resistance to these antibiotics was found in the current investigation. Morbidity and mortality rates are significantly affected by MDR bacterial strains. Organ transplantation, hip replacement surgery, chemotherapy for cancer treatment, and intensive care for preterm neonates are not often carried out in the absence of efficient antibiotics [27]. Human infections can result from a wide range of bacteria, including *S. aureus*. Most of these bacteria are known to exhibit specific virulence factors, which contribute to their pathogenicity. These virulence factors include the formation of biofilms, toxin synthesis, fimbriae, and pili. Of all these virulence mechanisms, the majority of recalcitrant infections are caused by biofilm formation, which is also the most difficult to treat because the organisms involved are highly resistant to antibiotics [10,11,22–25, 28]. The level of resistance to other studied antibiotics (erythromycin, co-trimoxazole, gentamicin, etc.) was lower in our study. These *S. aureus* strains responded well to vancomycin and linezolid. The MRSA prevalence in our study 68.7% is greater than what Al-Safani et al. [25] and Al-Akwa et al. [22] found for Yemen. However, it was comparable to previous worldwide research, where the range was between 40% and 69.8% [29–33]. In hospital settings, *Staphylococcus aureus*, or *S*. *aureus*, is commonly encountered. It adheres to host tissues and medical devices, where it continues to live. This could result in pneumonia, bacteremia, osteomyelitis, endocarditis, and infections of the skin and soft tissues [28, 34–36]. These infections are difficult to cure because of the biofilm produced that promotes the resistance of *S. aureus* to antibiotics [37]. Furthermore, the development of biofilms is thought to be a protected growth mode that allows bacteria to adapt to hostile settings [38]. In addition to shielding bacterial cells from harsh environments including intense heat, starvation, dehydration, and even antibacterial medications, the biofilm serves as a barrier to maintain a steady internal environment for bacterial cell activity [39]. As a result, bacteria are able to adhere better to the host surface, settle more quickly, defend themselves against the host immune system, and ultimately promote long-term infection. Thus, the first line of defense for bacteria is biofilm. Biofilm-forming bacteria are known to be resistant to the majority of antibiotics [40]. Planktonic bacterial cells are the target of most therapeutic antibiotic development. Planktonic cell-targeting antibiotics have the potential to exert selective pressure on bacteria, providing them with an edge over vulnerable competitors in terms of survival [41]. As a result, prolonged high-dose antibiotic therapy is typically necessary for antibiotic therapy against biofilm [42]. Chronic use of these antibiotics, however, may raise the risk of drug toxicity and antibiotic resistance [43]. Owing to the biofilm population’s high complexity and rapid adaptability [30], a thorough comprehension of the biofilm formation mechanism may offer new insights for the creation of efficient infection management plans that combat biofilms [31, 32].

*S*. *mutans* had 86.6% strains with moderate biofilm generation and 1.2% with strong biofilm production when compared to other *streptococci* in the *Streptococcus viridans* group (Table 2). Dental plaque is an oral biofilm that sticks to the teeth and is made up of several bacterial species, including *Streptococcus mutans*, which are embedded in salivary polymers and extracellular microbial products. Oral diseases result from the buildup of germs, which exposes the teeth and gingival tissues to high concentrations of bacterial metabolites [44]. Oxidative and acidic stresses commonly affect the biofilm that covers teeth [45,46]. Acid stress, which occurs when the pH in oral biofilms falls sharply to 4 or lower due to dietary carbohydrates, has been documented [46]. DNA becomes depurinated at 37°C, leaving apurinic (AP) sites [47], particularly in the case of guanine loss [48]. If dental plaque biofilm is allowed to grow over time, it may eventually lead to dental caries. When specific (cariogenic) microbiological populations start to predominate in an environment that supports them, there is an ecologic shift away from balanced populations within the tooth biofilm. The change to an acidogenic, aciduric, and cariogenic microbial community develops and is maintained by frequent consumption of fermentable dietary carbohydrate. A carious lesion, or cavity, is the symptom of this microbial shift in the biofilm, which is linked to an imbalance of demineralization over remineralization. This leads to progressive demineralization within the dental hard tissues (dentin and enamel first), with the activity shift causing acid production within the biofilm at the tooth surface. Dental caries can be prevented or arrested by stopping the dental plaque biofilm from developing or by restoring it to a non-cariogenic state [49,50]. Reducing the consumption of fermentable carbohydrates, or sugar, and regularly clearing the biofilm are two behavioral modifications that can help achieve this [49]. Though, in this investigation we noticed a high level of ampicillin resistance (96.3%) of *S*. *mutans*; and 19.5% for clindamycin. The current study demonstrated that *S. mutans* biofilms positively had a higher rate of resistance to tested antibiotics based on in vitro antibiotic sensitivity to different strains of the bacteria. These results may be attributed to the following facts: cells in biofilms become effectively resistant to drugs due to their modified metabolic activity and constitutive upregulation of drug pumps; additionally, *S. mutans* positive biofilms are resistant to standard antibiotics for Gram positive bacteria medications due to the availability of biofilms that are considered physical protection of *S. mutans* from medications [10,11,51].

## CONCLUSION

Our study demonstrated that 294 oral isolates (72.4%) formed moderate/strong biofilms, showing significant correlation with multidrug resistance (p <0.0001). *S. aureus* and *S. mutans* were predominant biofilm producers with antibiotic resistance rates of 95.1% and 96.3%respectively. These findings confirm biofilm formation as a major contributor of antimicrobial resistance in the oral cavity.

We recommend,

1. Clinical integration of biofilm susceptibility testing into clinical diagnostics,
2. Therapeutic development of anti-biofilm therapies for oral infections,
3. Establishment of stewardship programs to curb resistance evolution in Yemen.

This work directly addresses SDG-3 by combating a global health threat – antimicrobial resistance – through evidence-based strategies.

## DATA AVAILABILITY STATEMENT

All data generated or analyzed during this study are included in this published article (and its supplementary information files).

## ACKNOWLEDGMENTS

The authors would like to thank the Yemeni Ministry of Health and the Faculty of Dentistry at Sana’a University for their valuable support.

## CONFLICT OF INTEREST

The authors declare that there are no conflicts of interest.

## AUTHOR CONTRIBUTIONS

Nesreen Fadel Al-Sanabani: Conceptualization, Data curation, Formal analysis. Hassan Abdulwahab Al-Shamahy: Supervision, Validation, Writing – original draft.

## Notes

### Competing Interest Statement

The authors have declared no competing interest.

### Clinical Trial

N\A

### Clinical Protocols

N\A

### Funding Statement

The author(s) received no specific funding for this work.

### Author Declarations

This study was approved by the Medical Ethics and Research Committee of Sana'a University's Faculty of Medicine and Health Sciences (Approval No. 217, August 21, 2022). All participants provided written informed consent for oral swab collection.

